# Human studies of mitochondrial biology demonstrate an overall lack of binary sex differences: A multivariate meta-analysis

**DOI:** 10.1101/2021.11.03.21265871

**Authors:** Alex Junker, Jennifer Wang, Gilles Gouspillou, Johannes K. Ehinger, Eskil Elmér, Fredrik Sjövall, Kelsey Fisher-Wellman, P. Darrell Neufer, Anthony J.A. Molina, Luigi Ferrucci, Martin Picard

**Author notes:** Corresponding author 630 West 168th Street, Box 172 New York, NY 10032.

## Abstract

Mitochondria are maternally inherited organelles that play critical tissue-specific roles, including hormone synthesis and energy production, that influence development, health, and aging. However, whether mitochondria from women and men exhibit consistent biological differences remains unclear, representing a major gap in biomedical knowledge. This meta-analysis systematically examined 4 domains and 6 subdomains of mitochondrial biology (total 39 measures), including mitochondrial content, respiratory capacity, reactive oxygen species (ROS) production, morphometry, and mitochondrial DNA copy number. Standardized effect sizes (Hedge’s g) of sex differences were computed for each measure using data in 2,258 participants (51.5% women) from 50 studies. Only two measures demonstrated aggregate binary sex differences: higher mitochondrial content in women (g = 0.20, χ^2^ p = 0.01), and higher ROS production in skeletal muscle in men (g = 0.49, χ^2^ p < 0.0001). differences showed weak to Sex no correlation with age or BMI. Studies with small sample sizes tended to overestimate effect sizes (r = -0.17, p < 0.001), and sex differences varied by tissue examined. Our findings point to a wide variability of findings in the literature concerning possible binary sex differences in mitochondrial biology. Studies specifically designed to capture sex- and gender-related differences in mitochondrial biology are needed, including detailed considerations of physical activity and sex hormones.

## Introduction

Mitochondria are at the origin of multicellular life that gave rise to animal sexual reproduction, and to breathing, walking, and thinking *homo sapiens* (1). Mitochondria have shaped human physiology via energy transformation through oxidative phosphorylation (OxPhos) and via their multiple signaling functions (2). Over the last four decades, mitochondria have been implicated as a cause of rare (3) and common diseases including metabolic disorders, neurodegeneration, and cancer (4, 5). Mitochondria may also regulate the aging process in animals and humans (6–10), and serve as biomarkers of stress and disease (11, 12).

Consequently, the mitochondrion has become the most studied organelle in the biomedical sciences (13). This scientific focus on mitochondria emphasizes the need to understand why and how much mitochondria differ between individuals (inter-individual variation) among the diverse human population. Remarkably, as a field, we still do not know if, and to what extent, mitochondria display systematic binary sex differences between women and men. Resolving this question is paramount to developing clinically useful metrics of mitochondrial biology and to building comprehensive models of human health that incorporate bioenergetics.

Mitochondria are complex, multi-functional organelles. Within the cell, they have a life cycle (14), interact with one another through fusion processes (15, 16), and serve dozens of intracellular functions including ATP synthesis, reactive oxygen species (ROS) production, calcium handling, apoptotic signaling, redox homeostasis, hormone synthesis, and lipid and amino acid metabolism, among others (13). To accomplish specialized tasks in different cell types, mitochondria functionally specialize and acquire distinct tissue-specific proteomes and functional phenotypes (17, 18). For these reasons, single static molecular measures, such as mitochondrial DNA (mtDNA) copy number (mtDNAcn), provide little to no information about their functional capacity or complex biological phenotypes (19). Functionally meaningful mitochondrial metrics are best captured by dynamic measurements of functional capacity, such as respiratory capacity and efficiency (20), or ROS emission in the presence of specific metabolic substrates (21). Moreover, because mitochondria are multifunctional, mitochondrial phenotyping is most accurately achieved by probing in parallel multiple metrics of mitochondrial content and specific functions (e.g., (22)).

Recognizing the multiple roles and functions that mitochondria play in the organism highlights the inadequacy of the popular term “mitochondrial function.” By analogy, in human biomedical research, we do not speak of assessing “human function” – a category too imprecise to have empirical value – and instead rely on specific domains and subdomains of biology (locomotion, cognition, digestion). Similarly, our current understanding of mitochondrial biology makes it clear that speaking of “mitochondrial function” lacks the required specificity. Operationalizing different domains of mitochondrial biology to cover the major properties and unique functions of mitochondria provides a framework to systematically explore sex differences with the required degree of specificity (**Figure 1**).

**Figure 1.**
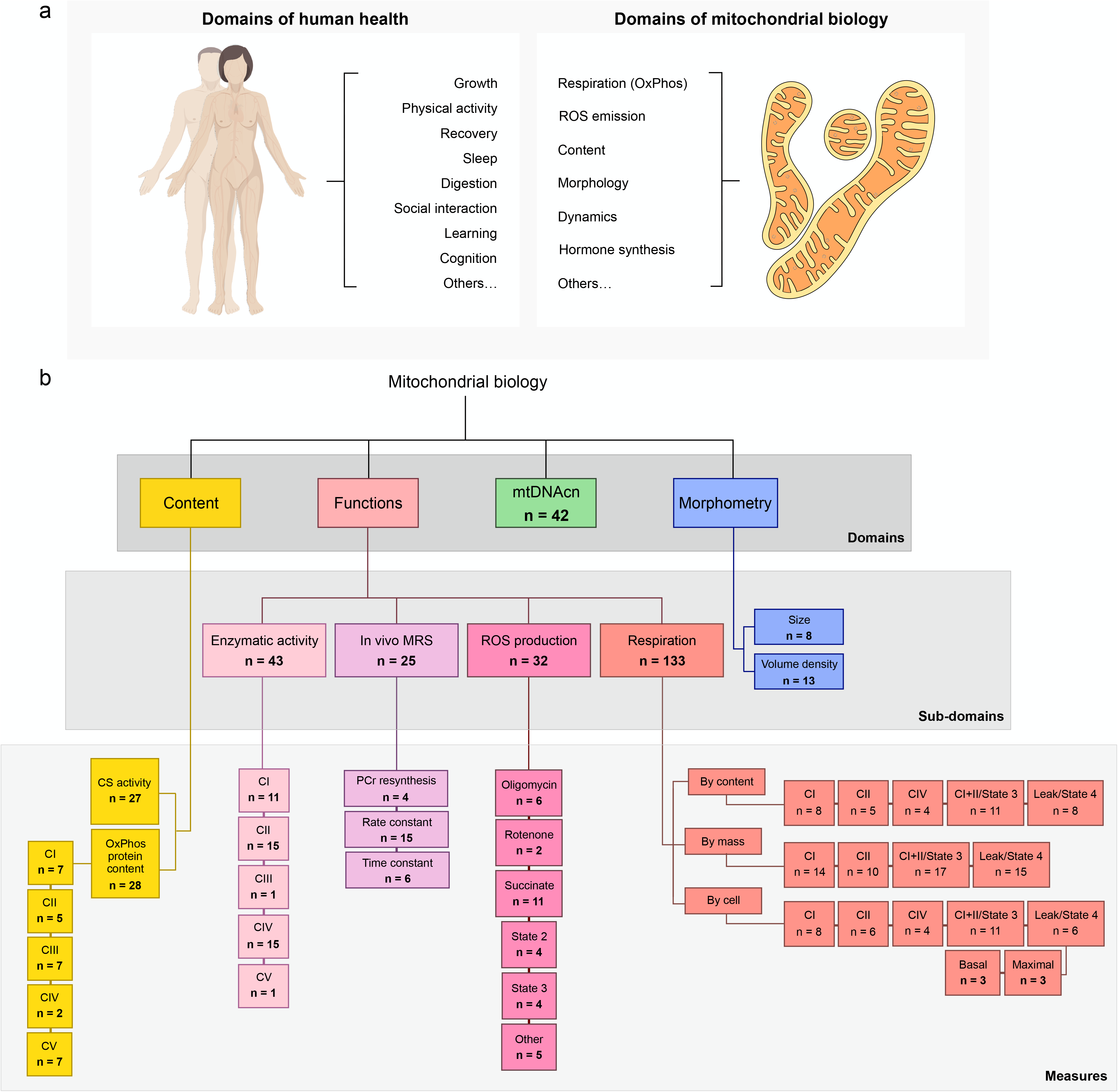
Mitochondrial biology encompasses a broad range of functions and characteristics. (**a**) Examples of biological domains observed in humans and mitochondria. (**b**) Mitochondrial perform several functions, and exhibit features that inform their overall phenotypes. Abbreviated taxonomy of mitochondrial biological domains examined in this paper. n = total number of measures across all included papers. CI – complex I, NADH dehydrogenase; CII – complex II, succinate dehydrogenase, CIII – complex III, ubiquinol-cytochrome c reductase; CIV – complex IV, cytochrome c oxidase; CV – complex V, ATP synthase; CS, citrate synthase; mtDNAcn, mitochondrial DNA copy number; MRS, magnetic resonance spectroscopy; PCr, phosphocreatine; ROS, reactive oxygen species

Three factors highlight the deep-rooted biological links between sex as a biological attribute and mitochondria. First, mammalian mitochondria are maternally inherited – meaning they are passed down solely through the oocyte (23). Second, an underappreciated fact is that the rate limiting step in the synthesis of all sex hormones, including estrogens, progestins, and testosterone (24) takes place in mitochondria, predominantly within the ovaries and testes. Indeed, the first enzymatic steps to the synthesis of *all* steroid hormones, which also includes glucocorticoids and mineralocorticoids, take place in the mitochondrial matrix (25). Third, mitochondria contain receptors for sex hormones. Both the estrogen receptor (Erβ) (26), and the androgen receptor (AR) (27) translocate to the mitochondrial matrix where they interact with mtDNA and influence multiple domains of mitochondrial biology. Thus, mitochondria contain the molecular machinery to i) produce and ii) sense the canonical hormones of sexual differentiation.

Previous studies have partially examined sex differences in mitochondria within different species, but we lack an overarching answer that considers the breadth of mitochondrial biology and of sexual characteristics in humans. In rats and mice, compared to males, mitochondria from females produce more energy and fewer ROS (28). However, it has been argued that the biological and social complexities of human gender/sex (29–31) make animal studies inadequate to explore sex differences in human mitochondrial biology. Human studies measuring sex differences in some aspects of mitochondrial biology have reported higher citrate synthase (CS, a marker of mitochondrial content) activity and greater antioxidant capacity in women’s mitochondria (28, 32), and comparable maximal respiration rates between binary sexes (33). Increasing evidence also suggests that sex hormones have a strong impact on mitochondrial biology independent of chromosomal sex and/or sex assigned at birth (34). Together, these findings highlight the need for a concerted community effort to systematically resolve the nature and magnitude of sex differences in mitochondrial biology among humans.

To address this need, we performed a quantitative, multivariate meta-analysis of available data on binary human sex differences, informed by a framework that incorporates multiple domains of human mitochondrial biology. When possible, we also explored whether methodological approaches (energetic substrates, normalization approach, isolated mitochondria vs. permeabilized cells/fibers methods) and individual traits (physical activity, age, and BMI) moderate the observed sex differences.

## Methods

### Outcomes

Mitochondrial biology was organized into four overarching domains: content, functions, mtDNAcn, and morphometry. Mitochondrial morphometry included two sub-domains, mitochondrial size and mitochondrial volume density. Mitochondrial content was assessed as CS activity and OxPhos complex protein content (CI - CV). Mitochondrial functions included four subdomains including: i) OxPhos enzymatic activity (CI - CV), ii) *in vivo* mitochondrial OxPhos capacity assessed through 31P-MRS, iii) ROS production, and iv) respiration. Mitochondrial respiratory capacity data was compared according to normalization method (normalized to mitochondrial content, tissue mass, or per cell), type of preparation (permeabilized fibers/cells, or isolated mitochondria), and respiratory state (state 3 phosphorylating respiration, or Leak/State 4 respiration). This organization is depicted in **Figure 1b**.

### Literature search

This meta-analysis was conducted according to the Preferred Reporting Strategies for Systematic Reviews and Meta-Analyses (PRISMA) guidelines (35). Authors J.W. and A.J. searched Medline via PubMed for eligible humans studies using two approaches: 1) a narrow search for studies that actively considered sex differences in mitochondrial biology, using the following search terms: mitochondria AND ("mitochondri* function" OR "functional capacity" OR respiration OR "ROS product*" OR "reactive oxygen species product*" OR "antioxidant activity" OR "ATP synthesis” OR “calcium uptake” OR “calcium handling” OR "electron transport chain" OR fusion OR fission OR “mitochondrial content”) AND ("men and women" OR "sex difference" OR "sex differences" OR "sexual dimorphism" OR "sex effects" OR "gender difference" OR "gender differences"); 2) a broad search for studies of mitochondrial biology, regardless of whether there was initial analysis by sex, using the following terms and filters: "mitochondrial function" NOT "review" ("muscle" OR "leukocytes" OR "brain"), using the filters Journal Articles, English, Human, Male, and Female. The last search was completed on July 17th, 2020. Investigators of included studies were also asked to recommend other relevant publications. Finally, the meta-analysis was advertised on social media and the community at large asked to suggest studies for consideration of inclusion.

### Inclusion/exclusion criteria

Criteria were established prior to searches and included only peer-reviewed articles, published in English, that examined at least one domain of mitochondrial biology (as defined above): mitochondrial content, OxPhos complex protein abundance or activity, in vivo magnetic resonance spectroscopy, respiration, ROS production, mtDNAcn, and mitochondrial volume or size. As calculations of standardized mean differences can be unreliable if the comparison groups sample sizes are severely imbalanced (36), studies had to contain no more than three times as many participants of one sex as another. Only studies of women and men without known illnesses (i.e., healthy controls) were included. Review articles and studies in animal models or cell culture were excluded. Studies of post-mortem tissues were included for their unique ability to enable mitochondrial measurements in otherwise inaccessible tissues such as the brain and cardiac muscle.

### Data extraction

The corresponding author(s) of each eligible article were contacted by email to request sex disaggregated data (mean, n, and SD), using a standardized template (**Supplemental File 1**). Follow-up emails were sent biweekly; after the third nonresponse, the first or senior author was also contacted. Returned data was entered into the data log, organized by domains of mitochondrial biology. Additional information was gathered for each study, including (where applicable) respiratory substrates used, methodological details (e.g., isolated mitochondria or permeabilized cells/fibers), average group age, BMI, and race, as available. The compiled data used in this meta-analysis is available as a resource in **Supplemental File 2** in the *Open Research* section of this article.

### Characteristics of included studies

Our searches yielded 1,212 publications in total. After screening abstracts, 275 full-text articles were evaluated for inclusion. Of these, 225 were excluded because they were either animal studies (n = 6), reviews (n = 7), lacked either women or men participants (n = 39), did not directly measure a functional or morphometric domain of mitochondrial biology (e.g., measured TFAM or UCP-1 protein abundance or gene expression) (n = 41), lacked healthy controls (n = 17), or had inadequate sample sizes (n = 98, see section “*Inclusion/exclusion criteria*”). An additional 17 studies were excluded because we failed to establish contact with the author, or sex-disaggregated data was not available. A detailed flowchart of the PRISMA-guided search process is available in **Figure 2**.

**Figure 2.**
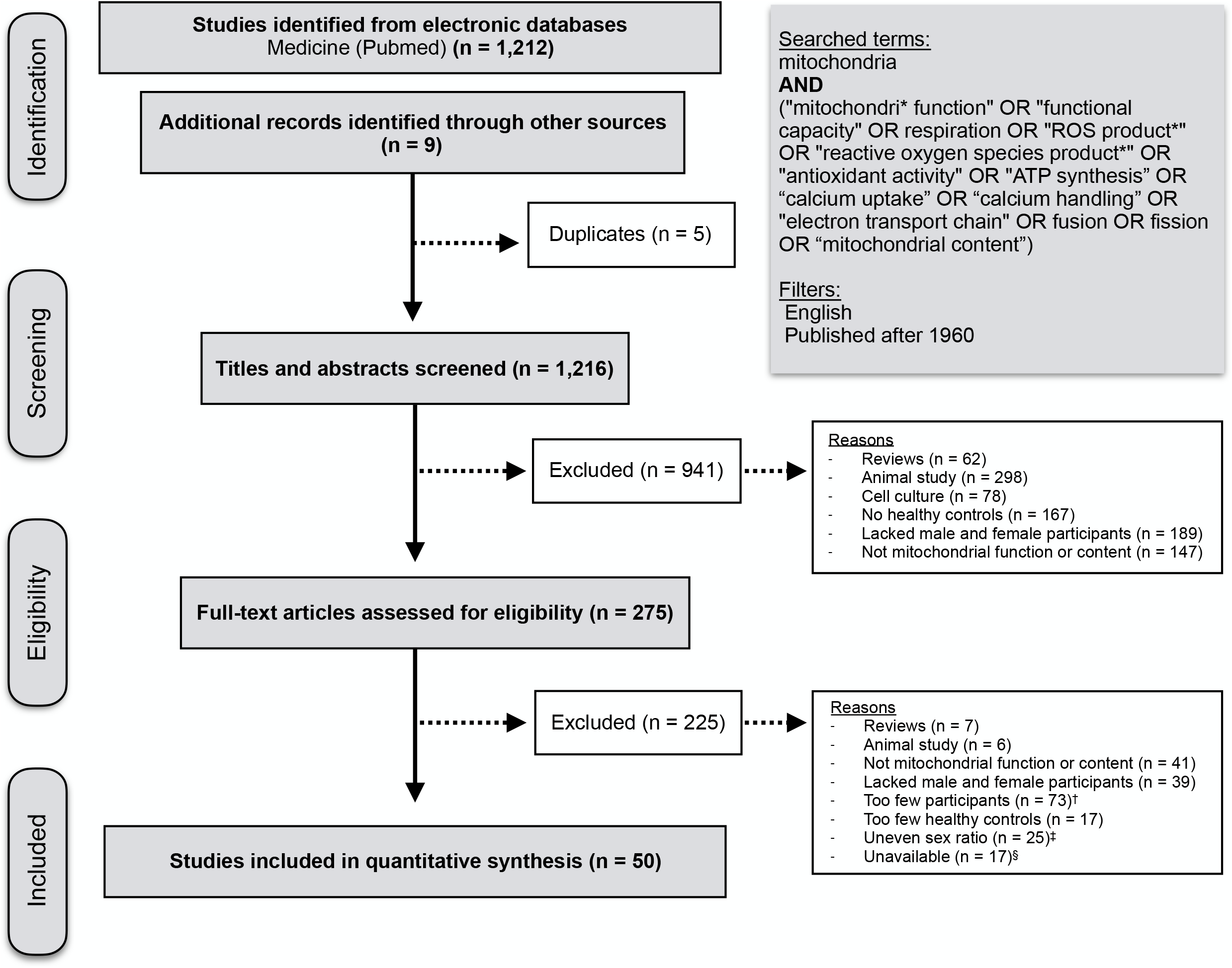
PRISMA flowchart of study selection process. Preferred reporting strategies for systematic reviews and meta-analyses (PRISMA) guideline flowchart of study eligibility and study selection. A full list of eligible articles is included in the first tab of Supplemental file 2. ^†^Studies were considered to have too few participants when the n for women or men was < 3. ^‡^Studies were considered to have an uneven sex ratio when there were over three times as many participants of one binary sex as the other (e.g., 30 men and 2 women). These studies were excluded because hugely unequal groups can produce unreliable effect size measures. ^§^Studies were considered unavailable when we 1) failed to establish contact with the author or 2) the author was not able to access sex-disaggregated data.

A total of 50 studies were included, resulting in a total of 2,268 participants (1,163 women and 1,105 men). Publication dates ranged from 1992 to 2021, with 23 studies (46%) published within the past 5 years. The average year of publication for included studies was 2013, compared to 2008 for unavailable studies, showing that data was more difficult to obtain from older studies. The average participants age ranged from 4 to 85 years (group mean), with almost half the studies (47%) with averages between 20-39 years. Average BMI was reported for 84% of studies and ranged from 22 to 34, with 55% of participant BMIs categorized as healthy under current guidelines (BMI < 25). 78% of studies did not report data on race or ethnicity. Of the six studies reporting racial data, three reported the race of each participant, and three reported solely the percentage of white participants. In these studies, the racial composition of participants ranged from 60-93% white, 0-30% Asian, 0-10% Black, and 0-5% Hispanic. No study reported gender/sex data beyond a binary sex designation (female or male). It is assumed that the vast majority of participants in this dataset are cisgender (i.e., their gender identity aligns with their sex assigned at birth), although this cannot be ascertained from the available data. This design limitation precludes interpretation of the relevance of these results for transgender and gender diverse (TGD) individuals (37). All studies reported gender/sex data as either a female or male designation, limiting analyses of sex only as a binary variable.

Physical activity directly influences several domains of mitochondrial biology (38, 39). Physical activity was an available variable in 5 studies: 4 comparisons of mitochondrial biology pre- vs. post-exercise interventions, and 2 comparisons of individuals with different activity levels (e.g., sedentary vs. active).

### Data synthesis and statistical analysis

PRISMA guidelines were consulted throughout the meta-analysis process to ensure standardized methodology. Effect sizes for each subdomain and individual measures were calculated as the standardized mean difference (Hedge’s *g*) between binary sexes for each measure of mitochondrial biology. A *g* value of ± 1.0 indicates that the two sexes differed by one standard deviation (S.D.). Hedge’s g was used as it includes a correction factor that outperforms Cohen’s *d* for small sample sizes (under n = 20) (36). As no field-specific effect size interpretation guidelines yet exist for mitochondrial biology, guidelines were adapted from a recent meta-analysis on metabolic syndrome components (40), whose cutoffs are comparable to those in other fields (41, 42). Effect sizes were considered negligible when falling between [absolute values] 0-0.19, small between 0.20-0.39, medium between 0.40-0.79, large between 0.8-0.99, and very large >1.00.

Forest plots show Hedge’s g with 95% confidence intervals (C.I.) with distinct background colors to indicate large effect sizes (g > +/- 0.8; yellow for higher in women, blue for higher in men). Results are organized by domain of mitochondrial biology, and secondarily by OxPhos complex, methodological approach, and specific measure, as appropriate. Effect sizes are displayed as calculated for all measures excluding MRS measures of phosphocreatine (PCr) time constant, for which the displayed effect size is the inverse of the calculated value (since time constant is considered roughly inverse to mitochondrial function), displaying the inverse of the measure allows higher values to be interpreted as ‘better’ function, aligning its interpretation with the rest of the forest plot.

For articles containing multiple reported measures with high biologic similarity (e.g., OxPhos CI+II and ETS CI+II measured in the same participants and tissues using the same methods), only one measure was included, selected based on methodological compatibility with other studies, which contributes to keep relative study weights comparable. As studies varied by more than two orders of magnitude in sample sizes, we chose to calculate an unweighted pooled estimate for each metric. Using this approach, each study has an equal weight on the overall estimate, rather than reflecting only the largest studies. Of the 50 original articles included, only 7 (14%) reported sex differences as a primary outcome, thus minimizing the risk of publication bias (i.e., studies showing significant sex differences would be preferentially published, and studies with non-significant ones would remain unpublished) for our primary outcome of sex differences.

For each sub-domain, the proportion of studies/measurements that were either ‘higher in women’ or ‘higher in men’ was compared using a χ^2^ tests of equal proportions where the null hypothesis is that an equal number of studies on either sides of the g = 0. All statistical tests were performed in Prism (version 8.4.3) or Excel (version 16.52), using two-sided distributions, with the level set to 0.05.

## Results

### Mitochondrial content is higher in women

Mitochondrial content was measured in 13 studies for a total of 27 measures (26 CS, one cardiolipin) across 264 participants (126 women) and 13 tissues (**Figure 3a**). Women had higher mitochondrial content in most studies (20 of 26, χ^2^ p = 0.01) for an overall estimate slightly higher in women (average g = 0.20).

**Figure 3.**
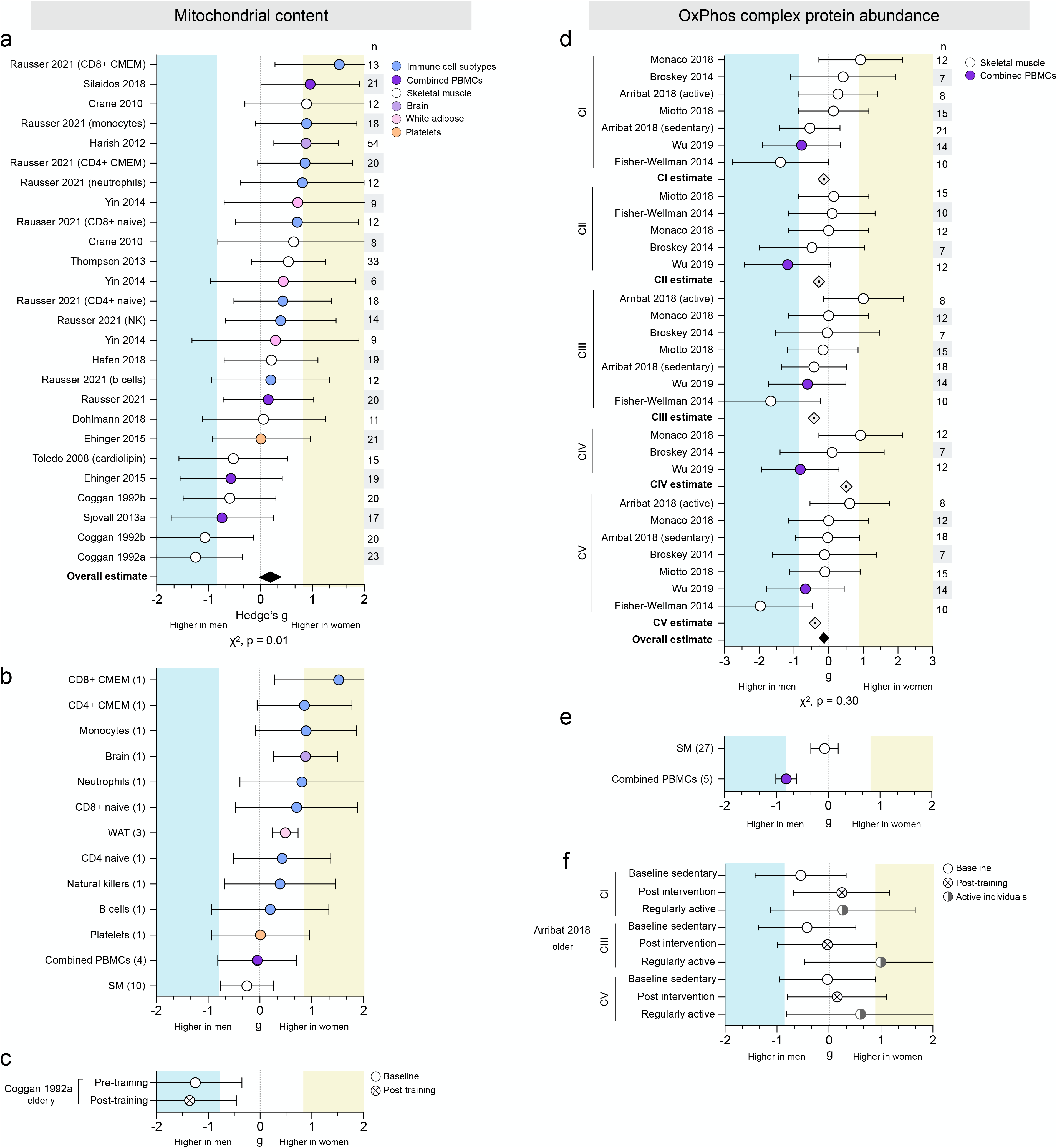
Mitochondrial content is consistently higher in women, but OxPhos complex protein abundance shows no consistent sex difference. (**a**) Forest plots of standardized mean difference (Hedge’s g) and pooled overall effect estimate (♦☐) for mitochondrial content (CS activity unless otherwise noted), color-coded by tissue. Values >0 indicate higher average production in women, and values <0 indicate higher production in men. Study n (women and men combined) is noted in the table to the right of the plot. (**b**) Tissue-specific average effect size for mitochondrial content; number of measures (papers combined) are noted on the left. (**c**) Standardized mean difference (Hedge’s g) and pooled effect estimate for CS activity before and after exercise training. (**d**) Hedge’s g, complex-specific average estimate (⍰), and pooled effect estimate (♦☐for oxidative phosphorylation (OxPhos) protein abundance, ordered by complex. (**e**) Tissue-specific average effect size for OxPhos protein abundance (all complexes combined). (**f**) OxPhos complex protein abundance in sedentary individuals (baseline and post exercise intervention) compared to regularly active individuals.

#### Tissue specificity

Mitochondrial content demonstrated potential tissue-specificity (**Figure 3b**). All isolated leukocyte cell types showed higher CS activity in women (from g = 0.20 to 1.52). Women also had higher CS activity in white adipose tissue (WAT) and brain tissue, (g = 0.49 and 0.88, respectively). However, each of these tissues (isolated leukocytes, WAT, and brain tissue) was measured in one study. Tissues measured across multiple studies, such as combined peripheral blood mononuclear cells (PBMCs) and skeletal muscle, demonstrated mixed sex effects by study and no overall sex differences (g = -0.05 and -0.20, respectively). Platelets were measured in one study and showed almost equal content in women and men (g = 0.01).

#### Exercise

One study assessing exercise in elderly individuals found that men had higher CS activity than women pre-exercise (g = -1.25, very large effect size), a difference marginally accentuated post-exercise (g = -1.37, very large) (**Figure 3c**).

### OxPhos protein abundance does not exhibit consistent sex difference

OxPhos complex protein abundance was measured in six studies for a total of 32 measures (CI, 8; CII, 5; CIII, 8; CIV, 3; CV, 8) across 87 people (42 women) and two tissues (**Figure 3d**). The measures were evenly distributed between higher in women or men for a negligible overall effect (average g = -0.19, χ^2^ p = 0.30). Most studies found higher CI abundance in women, but studies reporting higher CI abundance in men had larger effect sizes, resulting in a negligible CI-specific estimate (average g = -0.14). Similarly, measures of CII abundance were split between higher in women or men, for an overall estimate slightly higher in men (g = -0.28). A stronger effect was observed in both CIII and CV abundance, which were higher in men (g = -0.42 and g = -0.39, respectively). In contrast, CIV abundance was approximately equal in women and men (g = 0.07), indicating potential OxPhos complex-specific sex differences.

#### Tissue specificity

Skeletal muscle OxPhos abundance showed wide variation in both strength and direction of differences, for every OxPhos complex, resulting in a skeletal muscle-specific effect size of -0.27 (small, higher in men) (**Figure 3e**). In contrast, PBMC protein abundance was higher in men for all OxPhos complexes (from g = -0.61 to g = -1.18), resulting in a large leukocyte-specific effect size of -0.81 (higher in men). However, all PBMC measures came from one study, making it impossible to disentangle tissue-specific from method-specific factors.

#### Exercise

One study found higher protein abundance in active women than active men across all complexes (from g = 0.27 to g = 1.00) (**Figure 3f**). Sedentary men had higher protein abundance than sedentary women at baseline (from g = -0.03 to g = -0.48), but post-exercise intervention measures showed protein abundance was comparable between sexes (from g = -0.03 to g = 0.25).

### OxPhos enzyme activities do not exhibit consistent sex differences

Respiratory chain complex enzymatic activity was measured in 6 studies for a total of 40 measures (CI, 11; CII, 15; CIII, 1; CIV, 12; CV, 1) across 145 people (63 women) and 11 tissues (**Figure 4a**). The measures were evenly distributed between those higher in women or men and showed no overall effect (g = -0.01; χ^2^ p = 0.31). In analyses disaggregated by OxPhos complexes, CI activity measures showed no overall effect. Measures higher in women tended to demonstrate stronger effect sizes (up to g = 1.36), but the even distribution of values resulted in OxPhos complex-specific average effect sizes only negligibly higher in either women or men.

**Figure 4.**
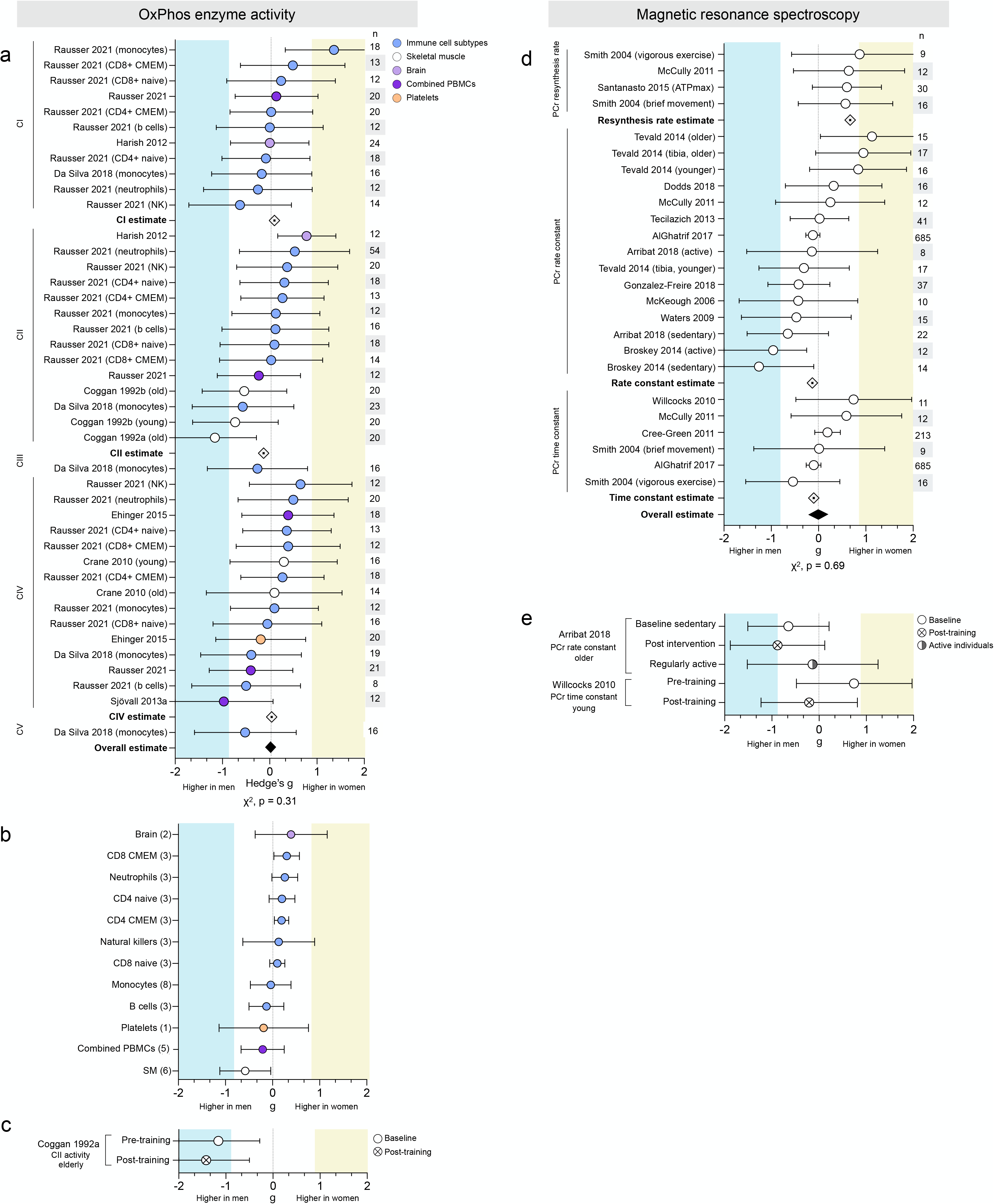
Neither oxidative phosphorylation enzyme activity nor in vivo MRS measures demonstrate consistent sex differences. (**a**) Forest plots of standardized mean difference (Hedge’s g), oxidative phosphorylation (OxPhos) complex-specific average estimate (⍰), and pooled overall effect estimate (♦☐for OxPhos enzymatic activity, ordered by complex and color coded by tissue. Values >0 indicate higher average production in women, and values <0 indicate higher production in men. Study n (women and men combined) is noted in the table to the right of the plot. (**b**) Tissue-specific average effect size for OxPhos enzyme activity; number of measures (papers combined) are noted on the left. (**c**) Hedge’s g for CII activity before and after exercise training. **(d)** Hedge’s g, measure-specific average estimate (⍰), and pooled effect estimate (♦☐for in vivo MRS, ordered by measurement. (**e**) In vivo MRS measures in sedentary individuals (baseline and post exercise intervention), compared to themselves and/or to regularly active individuals.

#### Tissue specificity

CII activity in isolated leukocyte subtypes was consistently higher activity in women (**Figure 4a**). Cell subtype data available from one study included both naïve and memory states of CD4+ and CD8+ T cells, two states well known to have different mitochondrial profiles (43). Tissue-specific average effect sizes concur, with six of the eight leukocyte subtypes showing higher activity in women (**Figure 4b**). In contrast, mixed PBMCs (a mixture of lymphocytes and monocytes) were only negligibly higher in men (g = -0.16). Skeletal muscle OxPhos enzyme activity was moderately higher in men (g = -0.58), while brain tissue was slightly higher in women (g = 0.39).

#### Exercise

One study of physical activity found that an exercise intervention tended to accentuate the higher skeletal muscle CII enzymatic activity (g = -1.16 and -1.42, respectively) (**Figure 4c**).

### Sex differences of in vivo mitochondrial OxPhos capacity may depend on the MRS metric used

In vivo phosphorus magnetic resonance spectroscopy (^31^P-MRS) was used to assess mitochondrial oxidative phosphorylation capacity in 14 studies for a total of 27 measures (PCr resynthesis rate, 4; PCr rate constant, 16; PCr time constant, 6) in skeletal muscle across 1,163 people (611 women) (**Figure 4c**). There was no data available for tissues other than skeletal muscle, or exercise-related analyses. Across all metrics, there is no evidence of sex differences (average g = 0.00, χ^2^ p = 0.78). Considered independently, both PCr rate constant and time constant showed negligible sex differences (average g = -0.13 and 0.10, respectively). In contrast, all measures of PCr resynthesis rate were higher in women ranging from medium to large effect sizes (g = 0.57 and 0.87, respectively), for a measure-specific average effect size of g = 0.67 (medium). Studies of PCr rate and time constant had significantly larger sample sizes (up to 685 people per measure) compared to studies of PCr resynthesis rate (n = 9 to 30 participants), which may contribute to the discrepancy in reported effect sizes.

### Mitochondrial ROS production is higher in men’s skeletal muscle

Mitochondrial ROS production was measured in 6 studies for a total of 31 measures across 152 people (90 women) (**Figure 5a**). There was an overall medium effect size showing greater ROS production in men than women (g = -0.49, χ^2^ p < 0.0001). The vast majority of measures were higher in men (88%), with effect sizes ranging from negligible (g = -0.03) to very large (g = -1.61). This effect was consistent across several experimental conditions, including in the presence of oligomycin (g = -0.47), rotenone (g = -0.07), succinate (g = -0.49), or other assorted carbohydrate or lipid substrates (g = -0.56).

**Figure 5.**
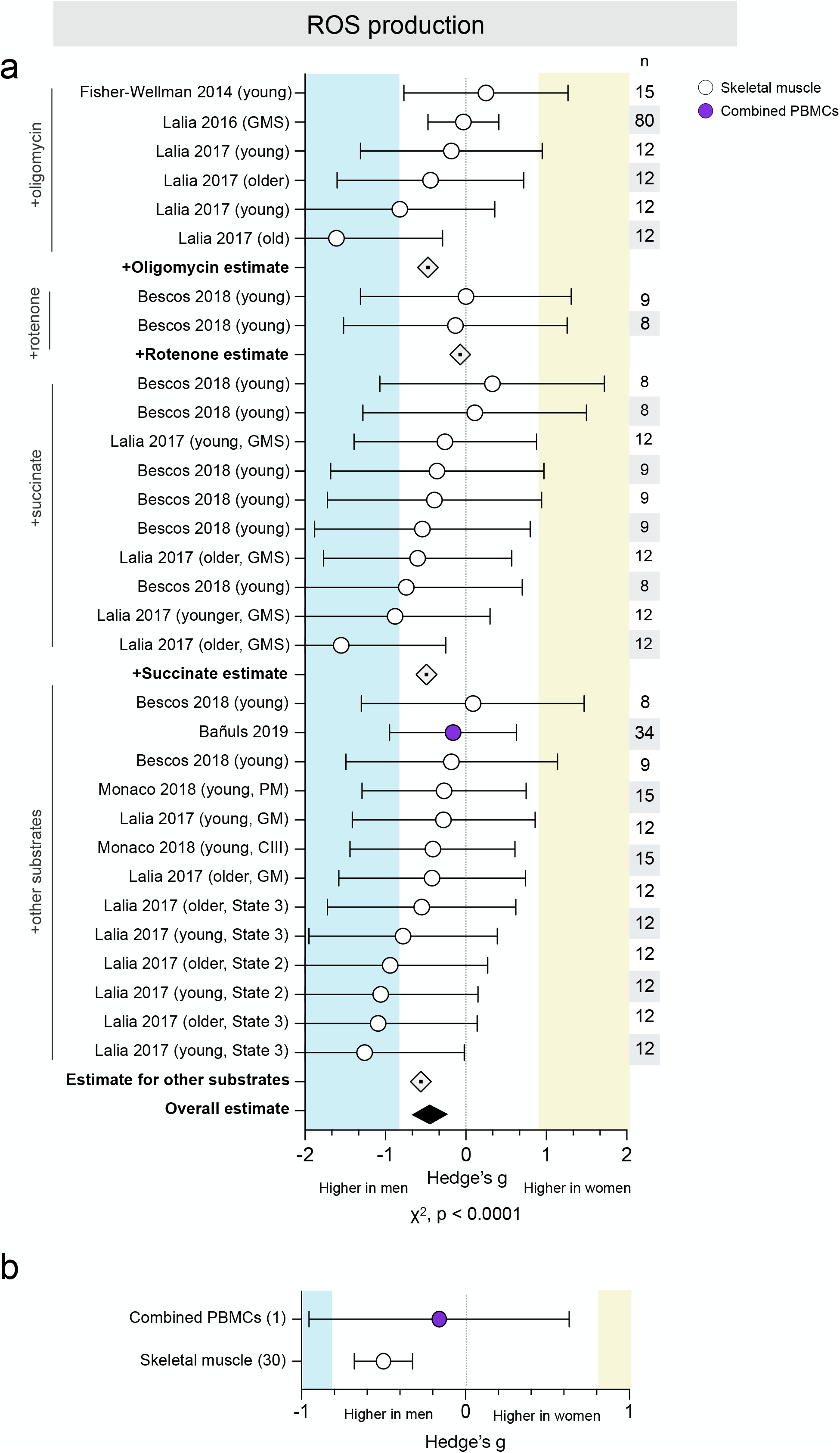
ROS production is consistently higher in men’s skeletal muscle. (**a**) Forest plot of standardized mean difference (Hedge’s g), substrate-specific average estimate (⍰), and pooled effect estimate (♦☐ for ROS production, color-coded by tissue and ordered by substrate (oligomycin, rotenone, succinate, and assorted carbohydrate and lipid). Values >0 indicate higher average production in women, and values <0 indicate higher production in men. Study n (women and men combined) is noted in the table to the right of the plot. (**b**) Tissue-specific averages for ROS production; number of measures (papers combined) are noted on the left.

#### Tissue specificity

Only one study measured ROS in combined PBMCs, finding negligibly higher production in men (g = -0.16) (**Figure 5b**). The remaining measures (30 of 31) were performed in skeletal muscle and yielded an average medium effect size (g = -0.50) indicating higher ROS production in men.

### Sex differences in phosphorylating respiration are sensitive to tissue and methodology

A total of 22 studies measured phosphorylating (state 3) respiration, utilizing a range of methodological and analytic approaches. Phosphorylating respiration was normalized to CS in 6 studies, resulting in 27 measures across 120 people (63 women) and three tissues (**Figure 6a**). The majority of these (27 of 37, 72%) were measured in permeabilized cells and myofibers and showed negligible sex differences in CI-stimulated (0.10), CII-stimulated (-0.07), and CI+II (-0.03) respiration, for no sex difference overall (average g = 0.06). The remaining 10 measures were evaluated in mitochondria isolated from skeletal muscle. Women had higher respiration for CI+II and CI-stimulated state 3 respiration (g = 0.26 and 0.56, respectively). In contrast, men had higher CII-driven respiration (g = -0.68), indicating that sex differences may be specific to OxPhos complexes. Overall, measures of respiration normalized to CS show no sex difference whether collected in permeabilized fibers or isolated mitochondria (g = 0.16), for overall no sex difference (g = 0.10, χ^2^ p = 0.31).

**Figure 6.**
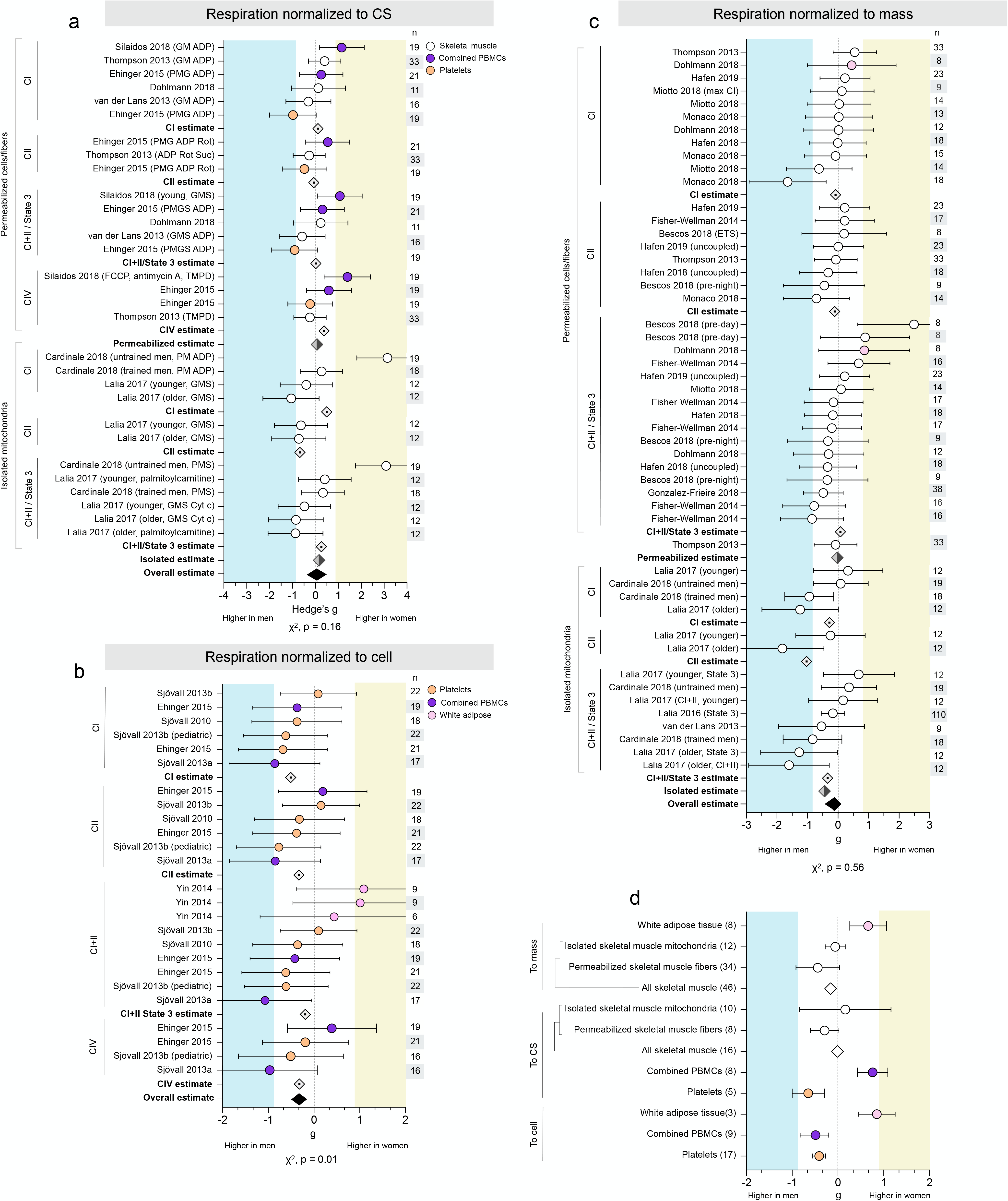
Sex differences in phosphorylating respiration are sensitive to tissue and methodology. Forest plots of standardized mean difference (Hedge’s g) for phosphorylating mitochondrial respiration normalized to (**a**) CS, (**b**) cell, or (**c**) mass, ordered by methodological approach (permeabilized cells/fibers vs. isolated mitochondria) and OxPhos complex, and color-coded by tissue. Including oxidative phosphorylation (OxPhos) complex-specific average estimates (⍰), method-specific estimates (⍰), and pooled overall effect estimate (♦☐). Values >0 indicate higher average production in women, and values <0 indicate higher production in men. Study n (women and men combined) is noted in the table to the right of the plot. (**d**) Tissue-specific average effect sizes for phosphorylating respiration, normalized to mass, CS, or cell. Number of measures noted to the left of the plot.

An additional 5 studies normalized phosphorylating respiration to cell count, resulting in 29 measures (CI, 8; CII, 6; CI+II, 11; CIV, 4; 26) from 87 participants (46 women) and three tissues (**Figure 6b**). Respiration was predominantly higher in men across all states, with average effect sizes ranging from negligible (g = -0.13, CI+II) to moderate (g = -0.51, CI), for an overall estimate slightly, but significantly, higher in men (g = -0.30, χ^2^ p = 0.01).

Phosphorylating respiration was normalized to tissue mass in 12 studies, resulting in 45 measures across 302 participants (161 women) and two tissues (**Figure 6c**). Most of these measures (36 of 45, 80%) were collected from respirometry performed in permeabilized myofibers or WAT cells and demonstrated no sex differences in CI-, CII-, or CI+II-stimulated respiration (g = -0.08, -0.11, and 0.08, respectively), for an overall absence of sex differences (average g = -0.02). The remaining 9 measures were from 3 studies that performed respirometry in mitochondria isolated from skeletal muscle. Average respiration was higher in men across all states, with effect sizes ranging from small to very large (CI, g = -0.28; CII, -1.03; CI+II, -0.34). Overall, phosphorylating respiration normalized to mass was moderately higher in men when measured in isolated mitochondria (g = -0.44), but not in permeabilized cells/fibers, for an overall lack of sex differences (g = -0.12, χ^2^ p = 0.56).

#### Tissue specificity

These measures of phosphorylating respiration demonstrated potential tissue-specific sex differences (**Figure 6d**). Regardless of methodological approach, women had higher phosphorylating respiration in WAT (g = 0.66, permeabilized cells; g = 0.85, isolated mitochondria). Platelet respiration was higher in men across all normalizations (g = -0.61, to CS; g = -0.41, to cell). However, sex differences in other tissues differed by method: women had higher respiration in PBMCs when normalized to CS (average g = 0.73), but men had higher respiration when normalized to cell count (average g = -0.49). Skeletal muscle respiration normalized to CS tended to be negligibly higher in women when measured in isolated mitochondria (average g = 0.16), but measures in permeabilized myofibers were higher in men (average g = -0.29, small effect size). In contrast, all measures of skeletal muscle respiration normalized to mass were higher in men (ranging from g = -0.06 to g = -0.44). When measures from both permeabilized fibers and isolated mitochondria are considered together, overall skeletal muscle respiration does not differ between binary sexes (from g = -0.01 to g = -0.16).

### Sex differences in mtDNAcn are tissue-specific

The number of mtDNA copies per cell, or mtDNAcn, was measured in 10 studies for a total of 42 measures across 308 people (146 women) and a wide range of tissues (**Figure 7a**). There were no consistent binary sex differences in mtDNAcn (average g = 0.01, χ^2^ p = 0.75) and, in contrast to other domains, there were 0 measures with strong sex effects for either women or men (g >/= 0.8).

**Figure 7.**
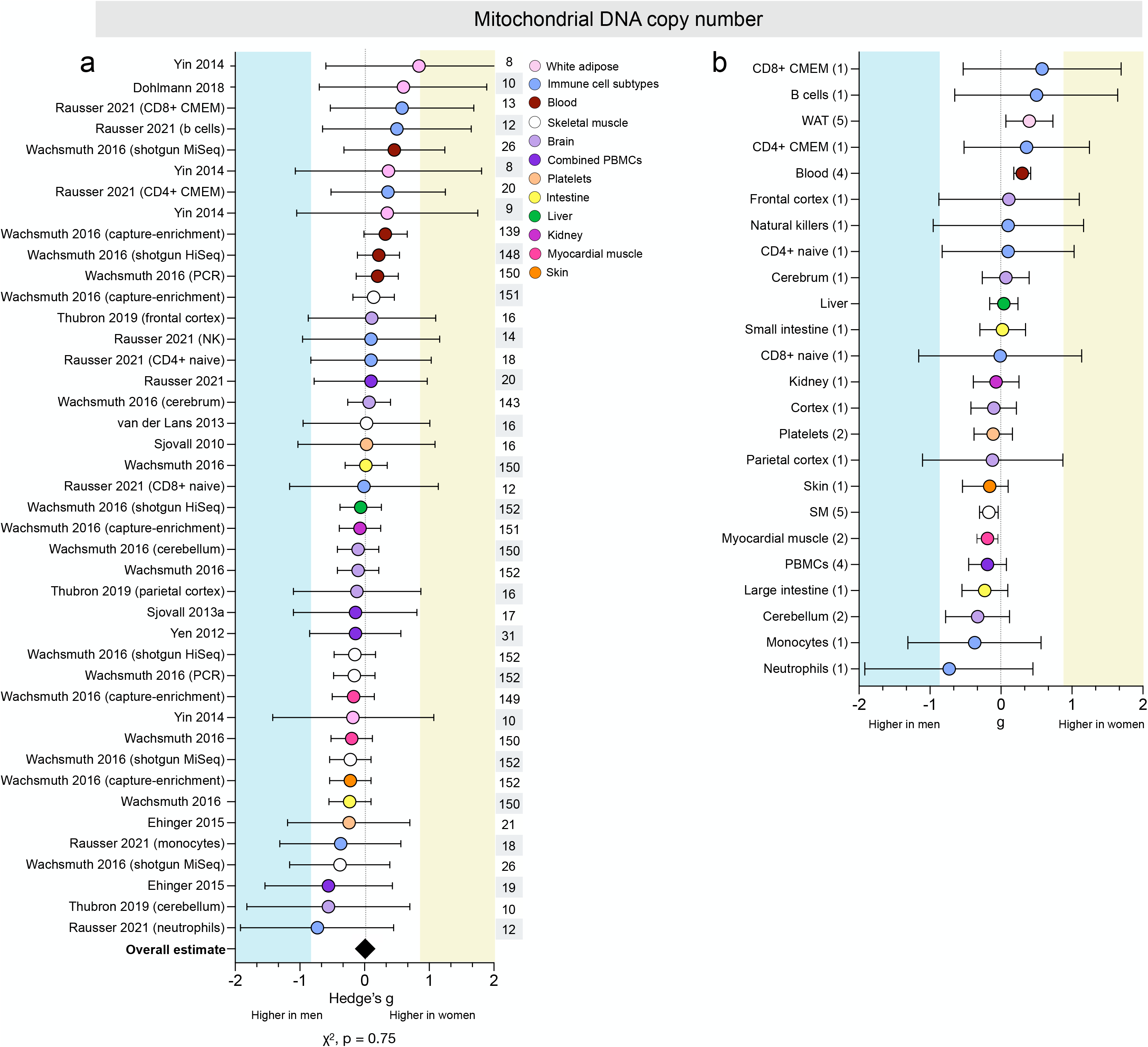
Sex differences in mtDNAcn are tissue-specific. (**a**) Forest plot of standardized mean difference (Hedge’s g) and pooled overall effect estimate (♦☐) for mitochondrial DNA copy number (mtDNAcn), color-coded by tissue. Values >0 indicate higher average production in women, and values <0 indicate higher production in men.Study n (women and men combined) is noted in the table to the right of the plot. (**b**) Tissue-specific average effect sizes for mtDNAcn; number of measures is noted in the table to the left of the plot.

#### Tissue specificity

Women had higher mtDNAcn across WAT and whole blood (g = 0.40 and 0.30, respectively) (**Figure 7b**). Several immune cell subtypes also showed higher mtDNAcn in women, including B cells (g = 0.50) and CD8+ & CD4+ CMEM cells (g = 0.58 and 0.36, respectively). In contrast, men had higher mtDNAcn for both neutrophils (g = -0.73) and monocytes (-0.37). However, all measures of mtDNAcn in immune cell subtypes came from one study, precluding interpretation on the effects of tissue-specific or method-specific factors.

### Skeletal muscle mitochondrial morphometry does not exhibit sex differences

Skeletal muscle mitochondrial volume density was measured in 7 studies for a total of 13 measures across 224 people (121 women) (**Figure 8a**). Only one measure showed substantially higher volume density in women (g = 1.15), while four measures demonstrated substantially higher volume density in men (ranging from g = -0.96 to -4.98). Overall, mitochondrial volume density appeared to be higher in men’s skeletal muscle (g = -0.60, χ^2^ p = 0.28). However, this difference may be exaggerated by the presence of an outlier with an effect size almost three times larger than any other measure of volume density (g = -4.98). When this measure is removed, the average overall mitochondrial volume density decreases by almost half, but remains higher in men (g = -0.31).

**Figure 8.**
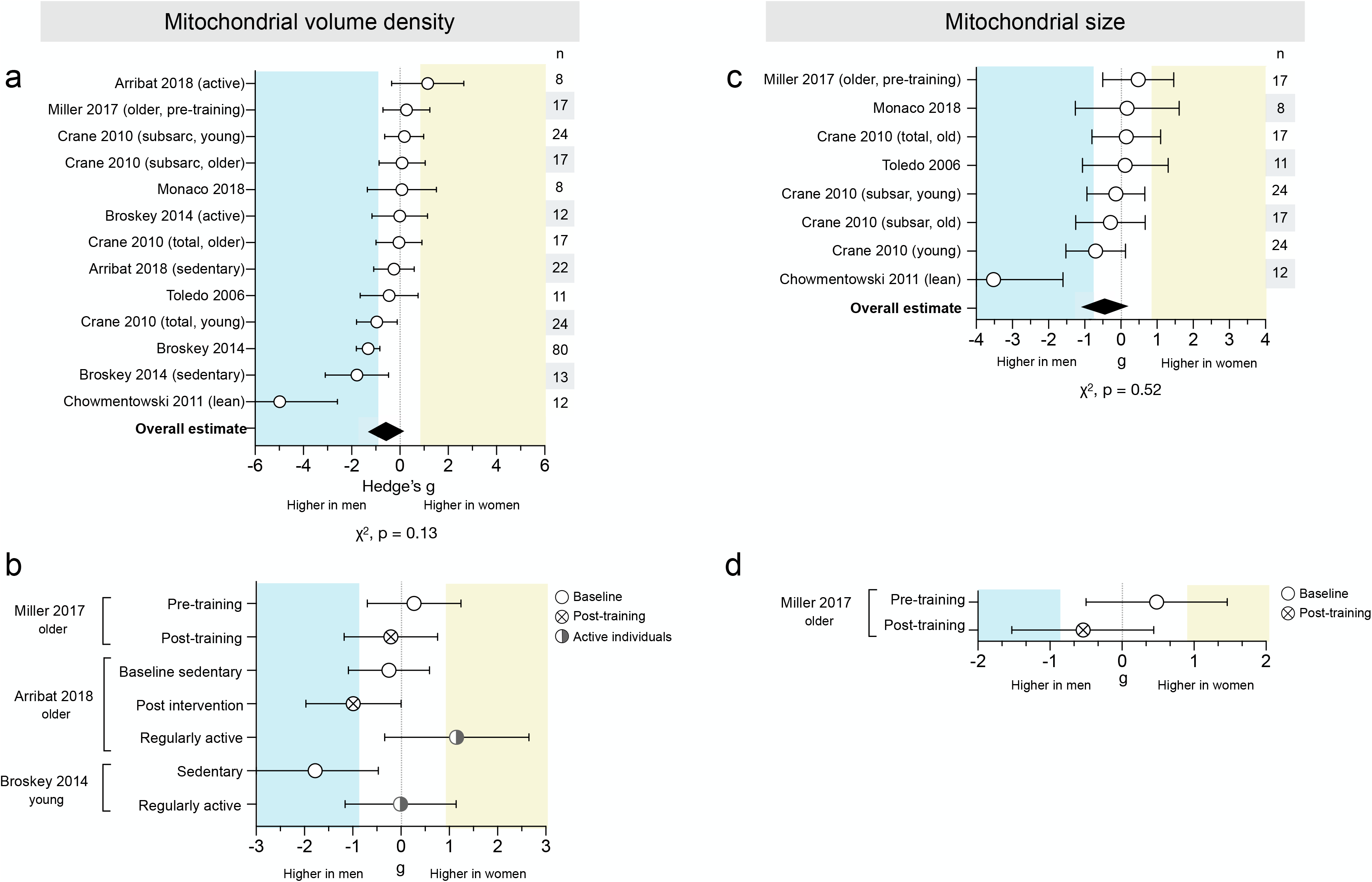
Skeletal muscle mitochondrial volume density and size lack consistent sex differences. (**a**) Forest plot of standardized mean difference (Hedge’s g) and pooled overall effect estimate (♦☐) for mitochondrial volume density. Values >0 indicate higher average production in women, and values <0 indicate higher production in men. Study n (women and men combined) is noted in the table to the right of the plot. (**b**) Measures of mitochondrial volume density in sedentary individuals (baseline and post exercise intervention), compared to themselves and/or to regularly active individuals. (**c**) Hedge’s g and pooled effect estimate (♦☐) for mitochondrial size. (**d**) (Measures of mitochondrial size in sedentary individuals (baseline and post exercise intervention)

Skeletal muscle mitochondrial size was measured using electron microscopy in 5 studies for a total of 8 measures across 89 people (49 women) (**Figure 8c**). The measures were evenly distributed between being higher in women or men (χ^2^ p = 0.52). The overall estimate was higher in men (g = -0.44), likely due to one measure with a very large effect size (g = -3.52). When this measure is removed, there is no sex difference in mitochondrial size (average g = -0.10).

#### Exercise

Two studies found higher volume density in men when assessed post-exercise intervention (g = -0.21 to -0.99) (**Figure 8b**), but pre-exercise measures varied, with one showing higher density in women (g = 0.27) and one showing higher density in men (g = -0.25). A third study found higher volume density in men among sedentary individuals (g = -1.78), but comparable density between sexes in active individuals (g = -0.01). One study assessed mitochondrial size in older individuals and found that women had larger mitochondria pre-training (g = 0.48), but men had larger size post-training (g = -0.54) (**Figure 8d**).

### Binary sex differences in mitochondrial biology show weak to no association with age and BMI

We systematically evaluated a potential moderating role of age and BMI on all domains of mitochondrial biology described above. Only two domains showed a statistically significant correlation with age: as a study group’s average age increased, i) ROS production became higher in men compared to women, and ii) respiration per cell changed from higher in men to showing no sex difference (**Figure 9a**). However, this dataset is limited in its ability to address this question. Most mitochondrial domains exhibited poor age resolution across the lifespan: for example, most studies of ROS production were conducted in individuals aged 20-30 or 70-80, making our analysis functionally more of a two-group comparison than a true correlation. This poor resolution, along with analyzing multiple tissues and methods together, makes it difficult to interpret whether sex differences in mitochondrial biology are moderated by age.

**Figure 9.**
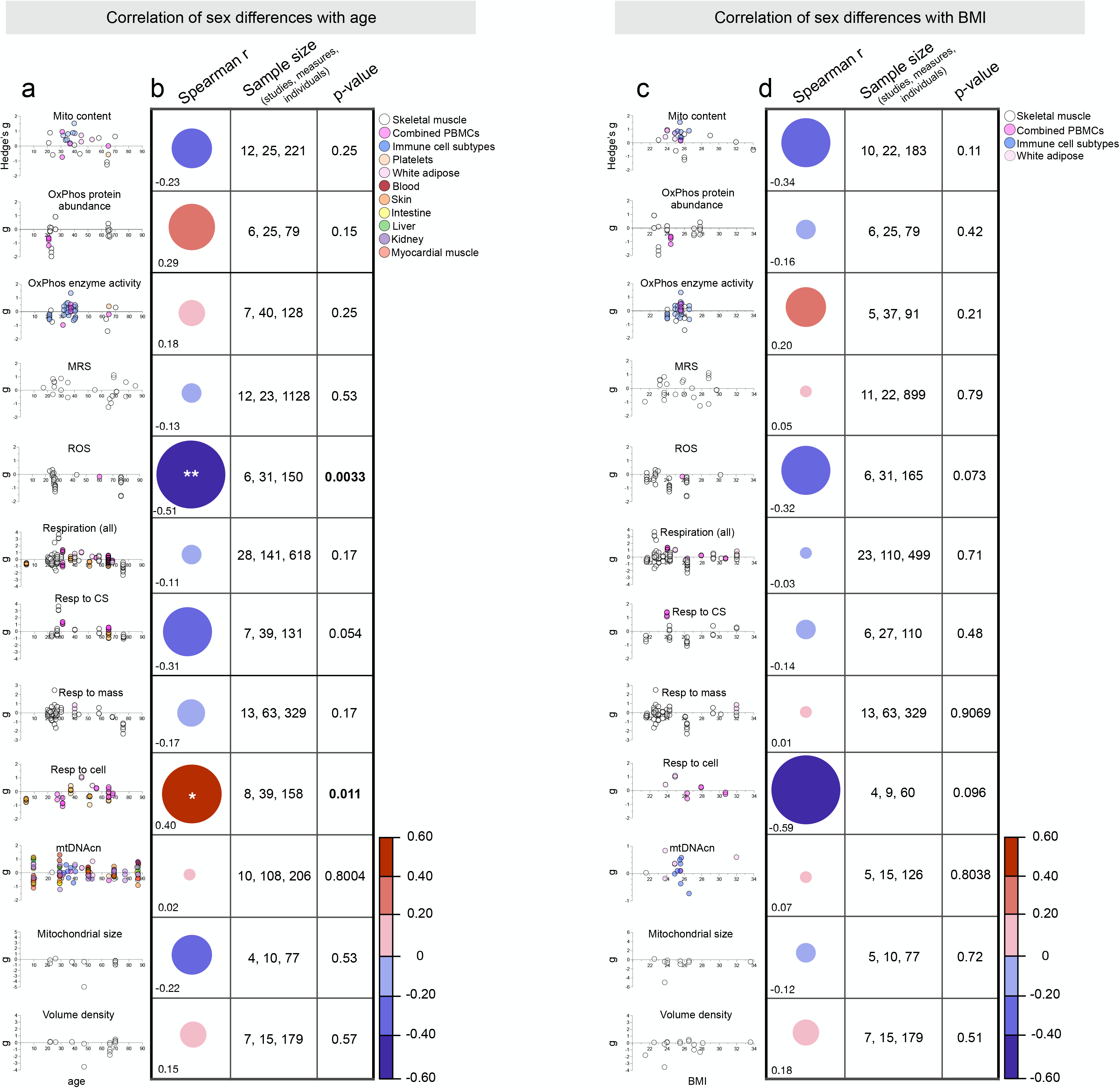
Binary sex differences in mitochondrial biology show weak to no association with age and BMI. (**a**) Correlation (Spearman r) between mitochondrial sex differences and average group age per measure, color-coded by tissue. The size and color of the circle represent the slope and strength of the correlation. Significantly significant correlations (p<0.05) are noted with an asterisk. Sample size is provided in terms of the number of studies, measures, and individuals. (**b**) Spearman r between mitochondrial sex differences and average BMI per measure, with points color-coded by tissue.

There were no significant correlations between mitochondrial biology and average group BMI, a limited and imperfect surrogate for body composition or obesity (44) (**Figure 9b**). The strongest negative correlation was for respiration to cell (r = -0.59, p = -0.096, suggesting that as BMI increases, the moderately higher respiration per cell in younger women approaches the null hypothesis. However, the relatively few measures on this analysis (9) provides poor resolution and little confidence in this conclusion. Near-continuous coverage in average BMI was only present for analysis of in vivo MRS and the combined respiration plot, boof which showed almost no correlation (r = 0.05 and -0.03, respectively).

### Influence of sample sizes on sex differences

To systematically assess the extent to which sample sizes may have influenced effect size estimates of sex differences across domains of mitochondrial biology, we plotted effect sizes as a function of sample size (**Figure 10a**). For each available measure, as expected (45), studies with smaller sample sizes yielded larger effect size estimates compared with larger studies particularly for three subdomains: MRS (r = -0.36, p-value = 0.04), mitochondrial respiration (r = -0.26, p-value = 0.006), and mtDNAcn (r = -0.38, p-value = 0.01). A similar association between sample size and effect size (r = -0.17, p-value < 0.001) was observed when all measures of mitochondrial biology analyzed together (**Figure 10b**).

**Figure 10.**
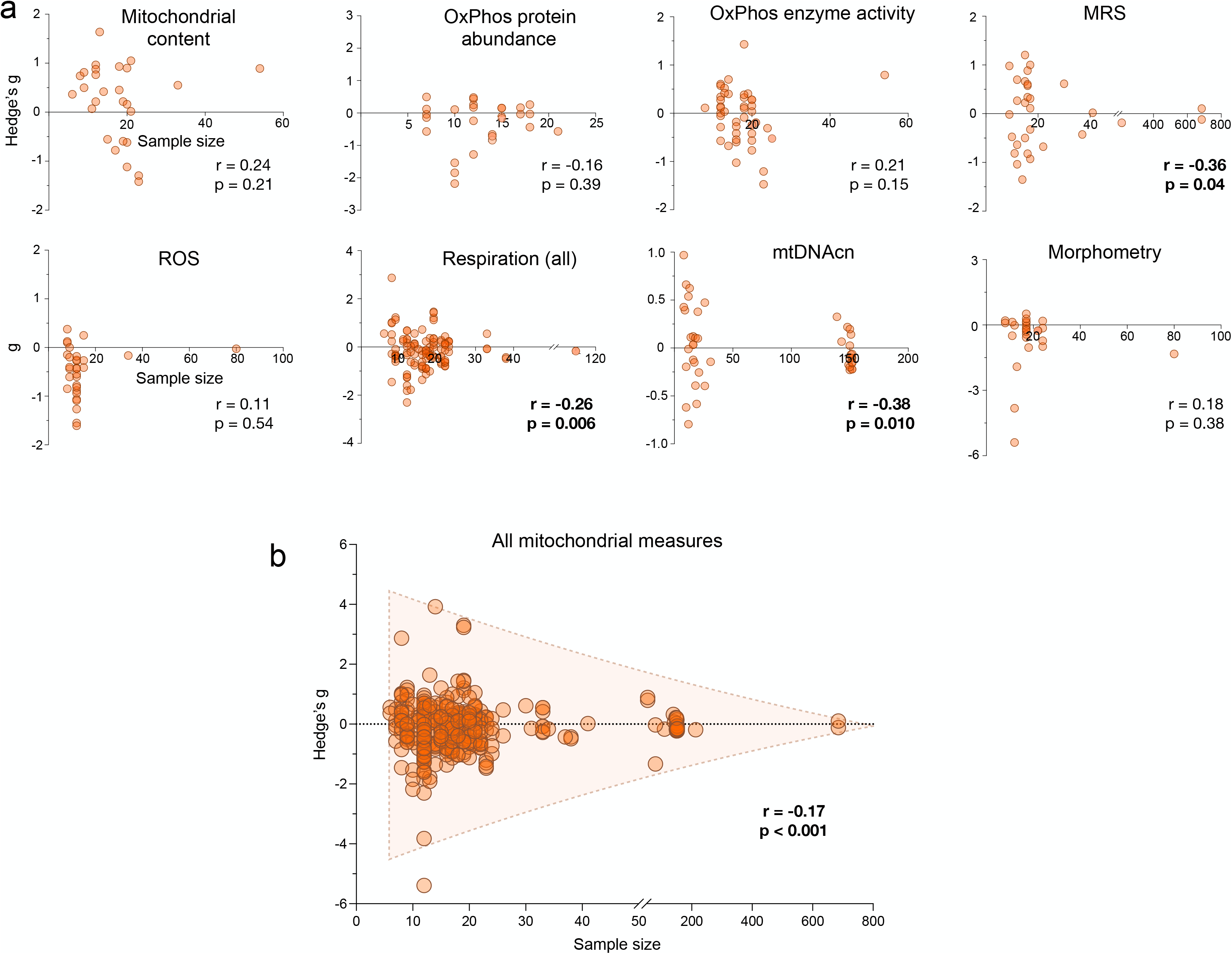
Observed sex differences in mitochondrial biology are negatively correlated with sample size. (a) Scatterplots showing the relationship between effect size and sample size by domain of mitochondrial biology. Each point is a measure from a study, previously represented by a row on a forest plot; measures >0 indicate higher values in women, and measures <0 indicate higher values in men. Presented correlations (Spearman r) and p-values were found by running correlations between the sample size and the absolute value of the effect size. (**b**) Scatterplot showing the relationship and Spearman’s r between sample size and effect size for all included measures.

## Discussion

In this meta-analysis of 39 distinct measures of mitochondrial biology from 50 studies combining >2,000 individuals, only two measures demonstrated consistent binary sex differences: higher mitochondrial content in women’s WAT and isolated leukocyte subpopulations, and higher ROS production in men’s skeletal muscle. Other measures showed inconsistent sex differences with large divergences in strength and direction across studies, experimental conditions (e.g., substrates), and sampled tissues. Sex differences across domains of mitochondrial biology showed weak to no interactions with BMI or age, although the available data offered sparse coverage of the human lifespan limiting the interpretation of these findings. Overall, these results indicate a lack of binary sex differences across most domains of mitochondrial biology, highlighting the need for adequately powered studies specifically designed to define the nature and magnitude of gender/sex and inter-individual differences in mitochondrial biology, and their relation to human health.

Three limitations of the current literature emphasize outstanding gaps in knowledge that poses challenges for the field. First, sex differences in mitochondrial health are heavily dependent on the tissue examined. For instance, women had higher respiration in WAT, while respiration appeared higher for men in platelets. This aligns with ongoing work characterizing biological sex variation within specific organ systems, such as the brain (46–48). Most available data included in this meta-analysis was derived from easily accessible tissues including blood, skeletal muscle, or adipose tissue; thus, whether less-studied tissues exhibit sex differences remains largely unknown. Sex differences may further vary between short-lived (certain blood leukocytes) and long-lived cells (neurons) due to their unequal cumulative exposure to sex hormones or other gendered influences. Second, little remains known about the interaction of sex and age on mitochondria. While the average ages among included studies ranged from 4 to 95 years, ∼50% of included studies mostly covered the third and fourth decades of life. Human aging involves well-documented changes in both sex hormone levels (49) and in mtDNA quality (50–52) but the relative paucity of data on older individuals in this analysis precludes definitive conclusions about whether sex differences are age-dependent. Additional work, particularly longitudinal studies, are therefore needed to characterize mitochondrial sex differences across the lifespan. Third, our results suggest that sex differences in mitochondrial biology may be sensitive to methodological (isolated mitochondria vs. permeabilized cells/fibers) and analytic (normalization to CS, cell, or mass) approaches. This sensitivity aligns with previous research showing that the effect of aging on mitochondrial respiratory capacity, ROS emission, and calcium retention capacity was exaggerated in isolated mitochondria compared to permeabilized myofibers (53), suggesting that inherent differences can be revealed or masked by different experimental preparations.

In designing future studies, the dynamic nature of mitochondrial biology should also be considered. For several psychophysiological phenotypes, within-person variation can be equal and up to four times larger than between-person differences shown at the group level (54). Thus, the biological significance of group differences, such as those meta-analyzed here, should be considered in light of how much natural variation can occur over time within individuals. Most available studies measured mitochondrial biology at a single time point; repeated-measures were only available in a small number of intervention studies (usually exercise) such that observed changes were attributed to the intervention’s effects. One proof-of-concept n-of-1 study documented 20 - 30% week-to-week variation in OxPhos enzymatic activities in sorted circulating leukocytes, suggesting substantial within-person variation in mitochondrial biology (22). We also previously found that up to 15% of the variance in PBMC mitochondrial OxPhos enzymatic activity normalized to mitochondrial content was predicted by psychological states the night before the blood draw (12), suggesting that mitochondria may dynamically adapt over short time frames and in response to more diverse factors than previously conceived. Overall, the natural variation in mitochondrial biology over time largely remains to be defined. Future studies specifically examining sex-related group differences in the context of intra-individual variation are needed to better understand the interplay of mitochondrial biology with different aspects of sex and gender.

Human sex is a complex system of biological features including, but *not* limited to, hormones, chromosomes, and tissues. Historically, these attributes have been reduced to a binary categorization of sex assigned at birth. Reducing this complexity into the binary “female or male” is a convenient categorization, but growing knowledge indicates that this binary categorization occludes our understanding of the true complexity of human sex and gender (often defined as socially-constructed roles and norms) (55–57). For example, one study following transmasculine people on gender-affirming hormone therapy (GHT) found that they had higher ROS production after three months taking testosterone (34). This finding suggests that variation in hormone levels – including between individuals with the same chromosomal sex – could influence mitochondrial behavior regardless of sex assigned at birth. Moreover, well-documented influences of gender as a social position on one’s diet (58–60), physical activity (61–64), sleep (65–67), and other health behaviors (68–70) are not captured by a binary sex classification, and may be responsible for physiological differences otherwise superficially attributed to biological sex (30). Sexual characteristics can also vary within a single person over time: gender role-related health behaviors change with marital status (71–73), and sex hormones are altered with parental role transitions (74), common conditions like polycystic ovary syndrome (PCOS) (75), and due to menopause or andropause (49, 76). On the one hand, it is possible that these additional factors simply add noise to the binary analyses reported above, thus reducing effect sizes. On the other hand, the influence of these factors on mitochondrial health could supersede that of sex assigned at birth, making the binary classification of limited use, possibly obscuring meaningful gender/sex-related variation. Research findings guided solely by a binary sex construct also are of limited applicability to TGD individuals, who represent a non-negligible and increasing fraction of the general population (37), and to intersex individuals (77, 78). A complete discussion of the limitations of binary sex constructs are beyond the scope of this meta-analysis but are explored elsewhere (29-31, 79, 80).

In summary, characterizing human sex differences in mitochondrial biology remains an important frontier and a collective challenge for biomedical scientists. While our meta-analysis reveals small-to-moderate binary sex differences among specific domains of mitochondrial biology, available evidence highlights the need for future adequately-powered, repeated-measures studies informed by a more nuanced and accurate understanding of gender/sex influences on mitochondria and cellular bioenergetics.

## Author contributions

J.W., A.J., and M.P. designed research. J.W. and A.J performed the literature review, collected, and organized data. A.J., G.G., J.E., and M.P. analyzed and interpreted data. A.J. prepared the figures. A.J. and M.P. drafted the manuscript. All authors edited and commented on the final version of the manuscript.

## Data Availability

All data for this study are openly available and can be accessed in Supplemental file 2.

## Acknowledgements

This meta-analysis was a group effort from investigators across multiple fields including metabolism/diabetes, exercise physiology, skeletal muscle biology, and gerontology. We are grateful to investigators and their team who provided sex-disaggregated data for this cross-disciplinary effort. Work of the authors was supported by FRQS grant 297877 to G.G., the Wharton Fund to A.J. and M.P., and NIH grants MH119336, AG066828, MH122706, and MH123927 to M.P.

## Nonstandard abbreviations

CI: complex I, NADH dehydrogenase
CII: complex II, succinate dehydrogenase
CIII: complex III, ubiquinol-cytochrome c reductase
CIV: complex IV, cytochrome c oxidase
CV: complex V, ATP synthase
CS: citrate synthase
GHT: gender-affirming hormone therapy
mtDNA: mitochondrial DNA
mtDNAcn: mitochondrial DNA copy number
MRS: magnetic resonance spectroscopy
OxPhos: Oxidative phosphorylation
PBMCs: peripheral blood mononuclear cells
ROS: reactive oxygen species
TGD: transgender and gender diverse

---

**Supplemental figure 1 – Leak/State 4 respiration does not demonstrate consistent binary sex differences.**

Forest plots of standardized mean difference (Hedge’s g) for nonphosphorylating mitochondrial respiration normalized to (**a**) CS, (**b**) cell, or (**c**) mass, ordered by methodological approach (permeabilized cells/fibers vs. isolated mitochondria) and OxPhos complex, and color-coded by tissue. Including oxidative phosphorylation (OxPhos) complex-specific average estimates (⍰), method-specific estimates (⍰), and pooled overall effect estimate (♦☐). Values >0 indicate higher average production in women, and values <0 indicate higher production in men. Study n (women and men combined) is noted in the table to the right of the plot. (**d**) Tissue-specific average effect sizes for phosphorylating respiration, normalized to mass, CS, or cell. Number of measures noted to the left of the plot. Nonphosphorylating respiration normalized to (**e**) CS or (**f**) mass in sedentary individuals (baseline and post exercise intervention). (**g** ) Hedges g and and pooled overall effect estimate (♦☐) for basal and maximal respiration in combined PBMCs.

**Supplemental file 1 – Data template**

**Supplemental file 2 – Data log**

