## Supplemental figure 1 for "Human studies of mitochondrial biology demonstrate an overall lack of binary sex differences: A multivariate meta-analysis"

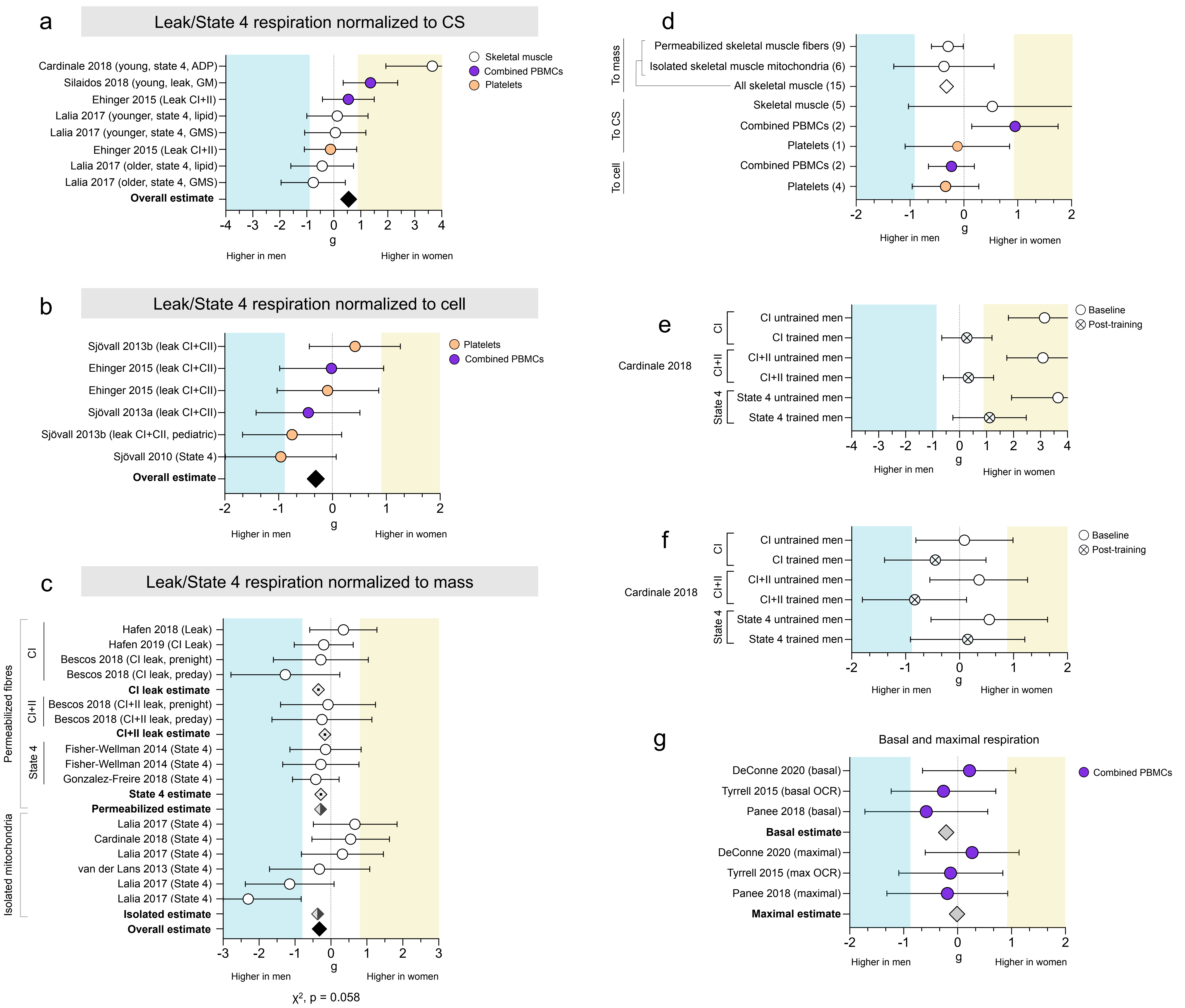


**Supplemental figure 1 – Leak/State 4 respiration does not demonstrate consistent binary sex differences.**
Forest plots of standardized mean difference (Hedge's g) for nonphosphorylating mitochondrial respiration normalized to (**a**) CS, (**b**) cell, or (**c**) mass, ordered by methodological approach (permeabilized cells/fibers vs. isolated mitochondria) and OxPhos complex, and color-coded by tissue. Including oxidative phosphorylation (OxPhos) complex-specific average estimates (◇), method-specific estimates (◆), and pooled overall effect estimate (♦). Values >0 indicate higher average production in women, and values <0 indicate higher production in men. Study n (women and men combined) is noted in the table to the right of the plot. (**d**) Tissue-specific average effect sizes for phosphorylating respiration, normalized to mass, CS, or cell. Number of measures noted to the left of the plot. Nonphosphorylating respiration normalized to (**e**) CS or (**f**) mass in sedentary individuals (baseline and post exercise intervention). (**g**) Hedges g and pooled overall effect estimate (♦) for basal and maximal respiration in combined PBMCs.
